# Left Atrial Appendage Closure in Patients with Atrial Fibrillation and Acute Ischemic Stroke Despite Anticoagulation

**DOI:** 10.1101/2023.08.15.23294148

**Authors:** Avia Abramovitz Fouks, Shadi Yaghi, Magdy Selim, Elif Gökçal, Alvin S. Das, Ofer Rotschild, Scott B Silverman, Aneesh B. Singhal, Sunil Kapur, Steven M. Greenberg, M. Edip Gurol

## Abstract

**Background:** The occurrence of acute ischemic stroke (AIS) while regularly using oral anticoagulants (OAC) is an increasingly recognized problem among nonvalvular atrial fibrillation (NVAF) patients. Due to the high risk of AIS recurrence (reported 5.3-8.9 per 100 patient-years) even when the type of OAC is changed, we aimed to elucidate the potential role of left atrial appendage closure (LAAC) for prevention of recurrent strokes among AIS-despite-OAC patients.

**Methods:** Through retrospective review of electronic medical records of a single hospital system between January 2015 and October 2021, we collected baseline and follow-up data from consecutive NVAF patients who had AIS despite regular use of OAC per current guidelines and subsequently underwent endovascular LAAC for recurrent stroke prevention. The primary outcome measure was the occurrence of AIS after LAAC, and the safety outcome was symptomatic intracerebral hemorrhage (ICH).

**Results:** Twenty-nine patients had the endovascular LAAC specifically because of AIS-despite-OAC. The mean age was 73.4 ± 8.7, 13 were female (44.82%). Mean CHA_2_DS_2_-VASc score was 5.96 ± 1.32, with an expected AIS risk of 8.44 per 100 patient-years. Fourteen of the patients had two or more past AIS-despite-OAC. After LAAC, 27 patients (93.10%) were discharged on anticoagulant which was discontinued in 17 (58.62%) after transesophageal echocardiogram (TEE) at 6 weeks. Over a mean of 1.75±1.0 years follow-up after LAAC, only one patient had an AIS (incidence rate [IR] 1.97 per 100 patient-years) and one patient had a small ICH (IR 1.97 per 100 patient-years).

**Conclusions:** LAAC in patients who had AIS-despite-OAC demonstrated a low annual rate of recurrent AIS in our cohort (1.97%) compared both to the expected IR based on their CHA_2_DS_2_-VASc scores (8.44%) and to the recent large series of AIS-despite-OAC patients treated with OAC/aspirin only (5.3%-8.9%). These findings support randomized trials of LAAC in patients who have AIS-despite-OAC.

## Introduction

Oral anticoagulants (OAC) are recommended by the American Heart Association guidelines for the prevention of cardioembolic stroke in patients with nonvalvular atrial fibrillation (NVAF).^1^ Compared to placebo, vitamin K antagonists (VKA) reduce the risk for acute ischemic stroke (AIS) and systemic embolism by 67%.^2^ Direct oral anticoagulants (DOAC) were found to be non-inferior to VKA for AIS prevention and proved to have lower intracerebral hemorrhage (ICH) risk, so DOACs are now adopted as first line drugs in many patients with NVAF. ^3-6^ However, OACs do not provide full protection from embolism even in low-risk patient populations and some patients still experience AIS while adequately taking their prescribed OAC. The risk of AIS in patients taking OAC is approximately 1.7% per year for VKA and 1.4% per year for non–VKA oral anticoagulants over 2.2 years follow-up in a population with mean CHA_2_DS_2_-VASc of 2.6. ^7^ The risk of having a recurrent AIS in patients who had an AIS-despite-OAC was 8.9 per 100 patient-years in a large multicenter study, about 7-10 folds higher than outcomes reported in DOAC arms of the randomized controlled trials of the 4 DOACs.^8,9^ Changing the type of OAC was not proven to affect the risk of recurrent stroke in this population.^9^ Therefore, patients who had an AIS-despite-OAC remain a challenging population for secondary stroke prevention. Left atrial appendage closure (LAAC) with the Watchman device (Boston Scientific Inc, Marlborough, MA) was approved by the FDA in 2015 and serves as an alternative stroke prevention method in patients with atrial fibrillation (AF) at increased risk of stroke who have a rationale to avoid long term anticoagulation.^10,11^ In this study, we aimed to elucidate the effects of LAAC on AIS recurrence among patients who had AIS-despite-OAC.

## Methods

### Patient selection

In this retrospective, observational study, we included consecutive NVAF patients who had endocardial LAAC with either Watchman 2.5/Watchman-FLX or Amplatzer Amulet (Abbott, Minneapolis, MN) devices within Massachusetts General Brigham (MGB) Health System (formerly known as Partners Health Care) specifically for AIS-despite-OAC, between January 2015 and October 2021. A thorough review of electronic medical records was performed by a neurologist and patients who were referred to LAAC specifically because of AIS(s) while adequately taking OAC as prescribed by their physician, were collected. All AIS on OAC happened while using DOAC regularly or while on warfarin with INR >2. Patients who were not using their OAC at the time of index AIS and patients who were not specifically referred because of AIS-despite-OAC were not included into this study.

### Standard Protocol Approvals, Registrations, and Patient Consents

This study was performed with the approval of and in accordance with the guidelines of the institutional review board (IRB) of MGB Health System. As this was a retrospective study, IRB waived the requirement for the informed consent. There were no photographs, videos or other information of any recognizable person.

### Outcomes

The primary outcome measure was the occurrence of symptomatic AIS during the follow–up period after LAAC. The safety outcome was the occurrence of symptomatic ICH during the same time period.

### Data collection and statistical analysis

Baseline data were collected including patient demographics and clinical characteristics at the time of LAAC. Cardiac imaging data before the LAAC, at time of procedure and at 6 weeks follow-up visit were collected. Periprocedural complications were defined as the occurrence of pericardial effusion/tamponade, vessel/cardiac perforation, device migration, major bleeding, stroke, death or any condition that required surgical or other intervention within 7 days of LAAC. Device-related thrombus and peri-device leak (including the size of the leak) were documented through review of cardiac imaging at all time points. Interval history from LAAC to last follow-up visit was reviewed for any event of AIS, symptomatic ICH including traumatic ICH, myocardial infarction, systemic embolism and major bleeding. Antithrombotic medication use was recorded at the time of discharge after LAAC, at 6 weeks after discharge, 3 months, 6 months, 12 months after discharge and during the last available follow-up visit. Detailed information related to all ischemic strokes before and after LAAC were registered. Clinical data as well as brain imaging, vascular imaging, cardiac imaging and lab results for each stroke were collected and reviewed. AIS pattern was determined based on diffusion weighed imaging (DWI) findings into the following subtypes: single lesion (cortico-subcortical lesion, cortical lesion, subcortical lesion ≥15mm, or subcortical lesion <15mm), two or more scattered lesions in one vascular territory, and multiple lesions in multiple vascular territories.^12^ Presumed cause of each AIS was concluded based on patient’s clinical characteristics, etiologic work-up and the imaging pattern of AIS on brain MRI, by consensus of a neurologist (AAF) and a stroke neurologist (MEG). For patients who had single subcortical infarcts of greatest diameter less than 15 mm, we also provided the location of the infarct. Such small infarcts might be more likely related to cerebral small vessel disease (cSVD) if they are located in classical deep locations such as internal capsule, basal ganglia, thalamus or pons whereas NVAF-related embolism might be the cause if they are in other subcortical locations such as corona radiata or centrum semiovale although cSVD remains in the differential in these patients as well.^13,14^ There were 3 patients who did not have a brain MRI available to review for all of their past strokes and the presence/absence of an embolic infarct was obtained from review of head CT and the official clinical/imaging reports. Categorical variables are reported as counts and corresponding percentage while continuous variables are reported as mean ± standard deviation (SD) or median (interquartile range [IQR]) depending on their distribution. Based on already published data, the baseline characteristics and the occurrence of AIS in follow-up of patients who did or did not have LAAC after AIS-despite-OAC were presented in 2 tables.

The report of the study follows the STROBE (Strengthening the Reporting of Observational Studies in Epidemiology) recommendations.

## Data Availability Statement

Anonymized data not published within this article will be made available by reasonable request from a qualified investigator.

## Results

Between January 2015 and October 2021, 29 patients were specifically referred for the LAAC procedure because they had AIS(s) while taking OAC as prescribed by their physicians. All patients were evaluated by stroke neurology physician. The mean age was 73.4 ± 8.7 years and 13 of the patients were female (44.82%). All patients had a diagnosis of NVAF (44.82% paroxysmal, 55.18% permanent) for a median duration of 4.83 years (IQR 1.52-10.11) at time of LAAC. Patients’ characteristics and vascular risk factors are further described in **Table 1**.

**Table 1.** Baseline patient characteristics of the study population (n=29)

|  |  |
| --- | --- |
| **Age, mean ± SD, years** | **73.4 ± 8.7** |
| **Female, n (%)** | 13 (44.8) |
| **AF, Paroxysmal, n (%)** | 13 (44.8) |
| **AF, Permanent, n (%)** | 16 (55.2) |
| **Hypertension, n (%)** | 28 (96.9) |
| **Diabetes mellitus, n (%)** | 15 (51.7) |
| **Dyslipidemia, n (%)** | 29 (100) |
| **CAD, n (%)** | 9 (31) |
| **PVD, n (%)** | 6 (20.7) |
| **DVT/PE, n (%)** | 4 (13.8) |
| **Heart failure, n (%)** | 10 (34.5) |
| **Chronic renal disease, n (%)** | 12 (41.4) |
| **Smoking, n (%)** | 1 (3.4) |
| **Past carotid endarterectomy/stenting, n (%)** | 3 (10.3) |
| **Multiple prior ischemic strokes, n (%)** | 14 (48.3) |
| 2 ischemic strokes | 11 (37.9) |
| 4 ischemic strokes | 2 (6.9) |
| 5 ischemic strokes | 1 (3.4) |
| **CHA<sub>2</sub>DS<sub>2</sub>-VASc, mean ± SD** | 5.96 ± 1.32 |
| **HAS-BLED, mean ± SD** | 4.24 ± 0.93 |
| **LAAC device type** |  |
| Watchman | 16 (55.2) |
| Watchman-FLX | 13 (44.8) |

SD, standard deviation; AF, atrial fibrillation; CAD, coronary artery disease; PVD, peripheral artery disease; DVT, deep vein thrombosis; PE, pulmonary embolism; LAAC, left atrial appendage closure.

Detailed characteristics pertaining to the patients’ AIS before LAAC as well as their management and outcome events during follow-up are provided under **Table S1**. Fourteen patients (48.3%) had more than one AIS prior to LAAC. Five patients had an AIS before LAAC while they were treated with VKA, 21 while treated with DOAC and 3 patients while they were treated with either VKA or DOAC at different time points as shown in **Table S1**. Upon detailed review of potential etiologies of AIS-despite-OAC, only one patient (**patient #16** in **Table S1**) had only one AIS consisting of a single small infarct in a classical deep location (thalamus) that could have been related to cerebral small vessel disease although embolism cannot be ruled out in the presence of NVAF. All other patients sustained clearly embolic looking infarcts while on OAC at least once, leading up to their referral to LAAC. No patient had proximal large vessel atherosclerotic disease or other classical cause for their AIS. The potential additional etiologic factors in 3 patients are provided in **Table S1**. Mean CHA_2_DS_2_-VASc score was 5.96 ± 1.32, with a calculated annual ischemic stroke risk of 8.44 per 100 patient-years. Mean HAS-BLED score was 4.24 ± 0.93. Sixteen patients (55.2%) had the Watchman 2.5 implanted while the other 13 patients had the new generation Watchman-FLX (44.8%). There were no peri-procedural complications for any of the patients. Patients were followed up for a mean of 1.75±1.0 years after LAAC. During follow-up, no patient had peri-device leak of more than 5 mm, however, 4 patients had peri-device leak of 3-5 mm. One of those patients had coiling of the leak with complete closure. None of the patients had device related thrombus. Post-LAAC antithrombotic treatment was individualized to the patients’ perceived needs and varied as described in **Table 2**. Twenty-seven patients were discharged on OAC. After the 6 weeks post-LAAC transesophageal echocardiogram (TEE) was performed, 12 (41.4%) patients remained on OAC treatment. At one year follow-up, one patient died and 2 were lost to follow-up. Among the remaining 26 patients, 18 patients were using antiplatelet monotherapy, one was not taking any antithrombotic treatment and 7 patients were still using OAC (24.1%).

**Table 2.** Antithrombotic treatment after left atrial appendage closure (n=29)

| **Antithrombotic treatment** | **Discharge** | **6 weeks p/LAAC** | **3 months p/LAAC** | **6 months p/LAAC** | **1 year p/LAAC** |
| --- | --- | --- | --- | --- | --- |
| **AP monotherapy** | 0 | 7 (24.1%) | 15 (51.7%) | 20 (69.0%) | 18 (62.2%) |
| **DAPT** | 2 (6.9%) | 10 (34.5%) | 4 (13.8%) | 0 | 0 |
| **VKA** | 1 (3.4%) | 0 | 0 | 0 | 0 |
| **VKA+AP** | 9 (31.0%) | 4 (13.8%) | 2 (6.9%) | 2 (6.9%) | 2 (6.9%) |
| **DOAC** | 5 (17.3%) | 0 | 0 | 0 | 1 (3.4%) |
| **DOAC+AP** | 12 (41.4%) | 8 (27.6%) | 7 (24.2%) | 5 (17.3%) | 4 (13.8%) |
| **None** | 0 | 0 | 0 | 1 (3.4%) | 1 (3.4%) |
| **Died** | 0 | 0 | 0 | 0 | 1 (3.4%) |
| **Lost to follow-up** | 0 | 0 | 1 (3.4%) | 1 (3.4%) | 2 (6.9%) |

p/LAAC, post left atrial appendage closure; AP, antiplatelet; DAPT, dual antiplatelet; VKA, vitamin K antagonist; DOAC, direct oral anticoagulation

For the primary outcome of recurrent symptomatic AIS, one patient had a small subcortical infarct in the centrum semiovale (< 15 mm) despite continued OAC use, 190 days after LAAC (patient #11 in **Table S1**). This patient had prior medical history significant for monoclonal gammopathy of unknown significance (MGUS) and 5 ischemic strokes prior to LAAC, 4 of those while taking DOAC. Accordingly, incidence rate (IR) for recurrent AIS after LAAC in our study population was 1.97 per 100 patient-years. For the safety outcome of symptomatic ICH, one patient had a small cerebellar ICH while taking DOAC and aspirin (IR 1.97 per 100 patient-years) 647 days after LAAC (patient #18 in **Table S1**). This patient had 4 ischemic strokes (2 while using DOAC, 2 while using VKA) leading up to the decision to perform LAAC, and a prior brain MRI also showing multiple mixed location (deep and lobar) cerebral microbleeds. There was no systemic embolism in any patient during follow up. None of the patients suffered myocardial infarction or major bleeding.

## Discussion

We performed a retrospective analysis of patients who had endocardial LAAC due to AIS-despite-OAC, in order to investigate the role of LAAC in this high-risk patient population. During a 1.75 years follow-up period, only one patient experienced an AIS after LAAC resulting in an IR of 1.97 per 100 patient-years. Our study did not have a control arm, but AIS IR was lower compared to the expected rate based on mean CHA_2_DS_2_-VASc (8.44 per 100 patient-years) and compared to previously published large series of patients who had AIS-despite-OAC and were kept on OAC without LAAC (5.3-8.9 per 100 patient-years) as shown in Figure 1. Outcome data from our consecutive case series support the view that LAAC might be a useful approach to decrease the risk of AIS in this high-risk population.

**Figure 1.**
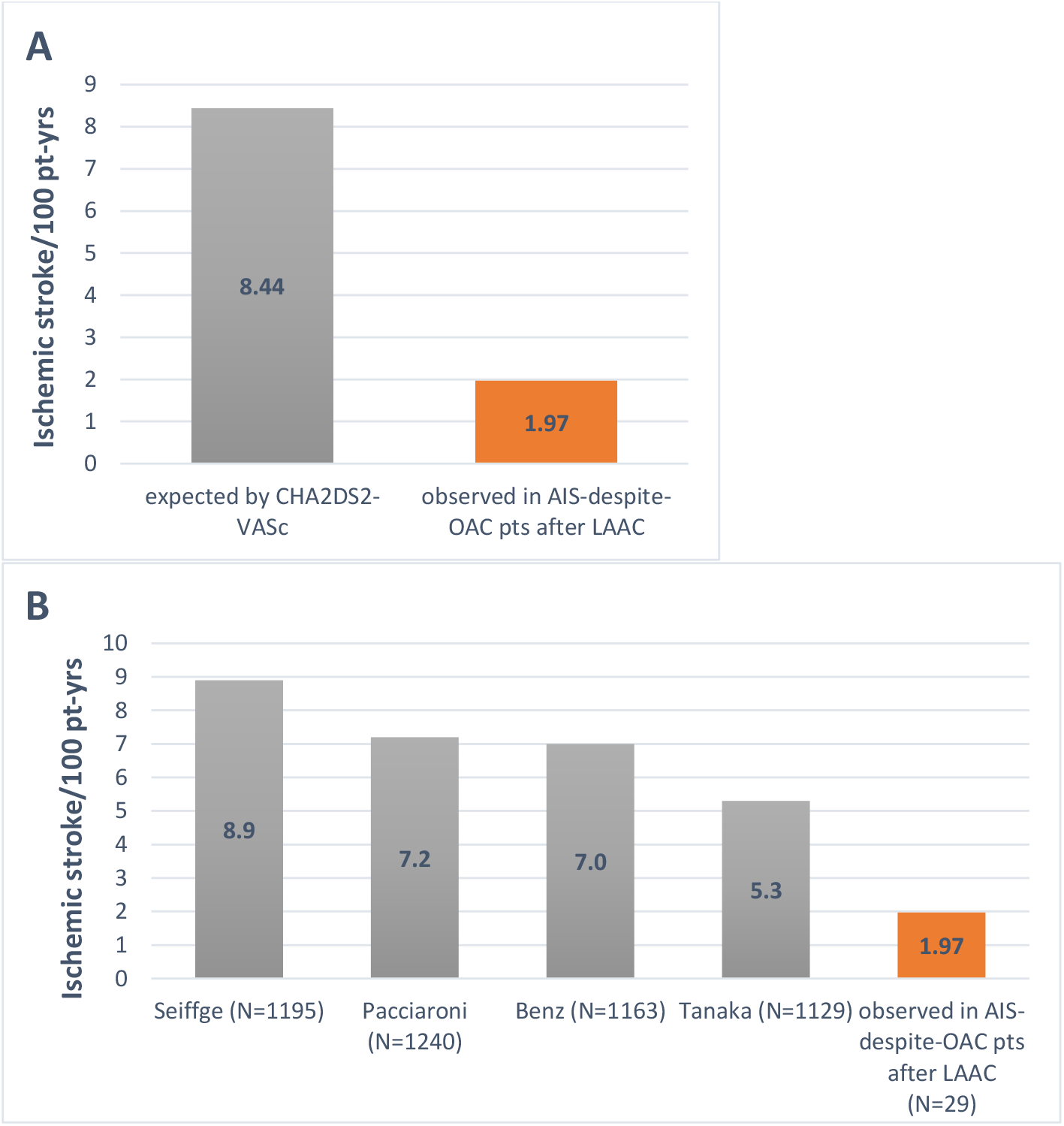
Recurrent ischemic stroke risk in patients with AIS-despite-OAC was lower after LAAC compared to expected by CHA_2_DS_2_-VASc (A) and compared to data from previous publications on AIS-despite-OAC patients who were kept on OAC as further described in Table 3 (B).

We report detailed information on index strokes, LAAC procedure and follow-up events within a well-defined patient population with NVAF who had LAAC specifically because they had one or more AIS-despite-OAC. NVAF patients with AIS-despite-OAC pose a therapeutic dilemma regarding the best secondary stroke prevention method. Multiple studies focused on longitudinal follow-up of large NVAF patient populations who had AIS-despite-OAC have been recently published (**Table 3**). ^9,15-20^ These patients were all kept on OAC (change in type/brand allowed) with or without addition of antiplatelet. These studies consistently showed high AIS recurrence rates ranging between 5.3-8.9 per 100 patient-years. Changing the type of OAC, the OAC brand or adding aspirin did not reduce the risk of recurrent AIS.^9^ Our study included a high embolic risk NVAF population with half of our patients having sustained more than one AIS-despite-OAC. Despite the high embolic risk patient population included (mean CHA_2_DS_2_-VASc: 5.96), the IR of 1.97 per 100 patient-years represents a 77% relative reduction in AIS risk based on the expected annual AIS rate in a patient population with similar mean risk score (8.44 per 100 patient-years). LAAC is not commonly performed in AIS-despite-OAC hence our relatively small study population, but we have been able to report very detailed data on the index strokes and follow-up thanks to the design of our study that was performed in a single hospital system composed of multiple hospitals. RCTs are needed to evaluate whether LAAC is superior to simple OAC continuation in patients with AIS-despite OAC. Based on a conservative 2-years cumulative IR of ischemic stroke of 10.3% in a NVAF patient population who had AIS-despite-OAC and kept on OAC, and on an IR of 4% in patients who had LAAC after AIS-despite-OAC based on our findings, an RCT including 698 patients would have 90% power to show superiority of LAAC over OAC-only approach.^15^

**Table 3.** Published data on baseline characteristics and outcomes in patients who had AIS-despite-OAC and kept on OAC without left atrial appendage closure.

| Author, year | Study design | No. of pts. | Age (years) | CHA<sub>2</sub>DS<sub>2</sub>-VASc | HAS-BLED | Follow-up | Recurrent AIS risk | Recurrent ICH risk |
| --- | --- | --- | --- | --- | --- | --- | --- | --- |
| Seiffge, 2020<sup>9</sup> | Pooled analysis from prospective, observational registries | 1195 | 79 (73–84) | 5 (4–6) | 3 (3–4) | 318 days | 8.9 per 100 patient-years | 2.0 per 100 patient-years |
| Benz, 2023<sup>15</sup> | Individual participant data from five randomized trials | 1163 | 73 (67–78) | 4 (3–6) | N/A | 337 days | 7 per 100 patient-years | N/A |
| Tanaka, 2020<sup>16</sup> | Pooled analysis from prospective, observational registries | 1129 | 75 (70–80) | N/A | N/A | 1 year | 5.3 per 100 patient-years | 0.6 per 100 patient-years |
| Pacciaroni, 2022<sup>18</sup> | Prospective, Observational | 1240 | 78.9 ± 9.1 | 77.1% ≥4 | 38.1% ≥ 3 | 15 months | 7.2 per 100 patient-years | 1.5 per 100 patient-years |

AIS, acute ischemic stroke; OAC, oral anticoagulant; ICH, intracerebral hemorrhage

Although it is difficult to confirm that an AIS is directly related to embolism from NVAF, the review of the stroke imaging for location/size/pattern of infarct(s) and ruling out alternative etiologies increase our confidence in the stroke etiology. Such review is difficult in large scale studies including randomized controlled trials (RCT) because of the bulk of the data that needs to be obtained and reviewed. Thanks to the design of our study based on a single healthcare system, we have been able to review and report clinical details as well as imaging data of 50 AIS that our 29 patients sustained before LAAC as well as the one AIS after LAAC. There was only one patient who suffered from one subcortical infarct less than 15 mm in diameter prior to LAAC, whereas the other patients had at least one clearly embolic infarct. The location of the small infarct in this patient was thalamus, which could have been due to either cerebral small vessel disease or NVAF-related embolism. Despite having very high embolic risk (high CHA_2_DS_2_-VASc scores and past AIS-despite-OAC), our cohort exhibited low rates of AIS recurrence after LAAC, similar to recent studies that explored the role of LAAC in patients with AIS-despite-OAC (**Table 3**). With respect to the etiology of the only one recurrent AIS after LAAC in our cohort, it is possible that the patient’s pre-existing MGUS might have played a role as a competing etiology for the patient’s pre-LAAC ischemic strokes as well as post-LAAC stroke.^21^ It is indeed important to perform a thorough stroke etiologic investigation among NVAF patients who had AIS-despite-OAC but there are many situations in which the relative contribution of a potential etiologic factor is unknown. If such a potential additional etiology is found, specific treatment can be planned, if available, in a hope to reduce the risk of further strokes. Etiologic evaluation and treatment decisions require a multi-disciplinary approach based on the complexity of the patient’s other medical conditions, as it was the case for patients enrolled in this study. All patients were evaluated by stroke neurology and cardiology specialists. Our findings suggest that in NVAF patients with AIS-despite-OAC mainly without a concurrent etiology, LAAC is associated with a low risk of recurrent embolic events in follow-up.

Current FDA approval allows the use of either VKA or DOAC or dual antiplatelets during the first 6-weeks immediately following LAAC with Watchman 2.5 or Watchman-FLX devices.^1^ The great majority of patients are taken off of anticoagulant therapy after the first follow-up TEE at 6 weeks, provided that there is no significant peri-device leak or device-related thrombus. Again, based on current FDA-approval, most patients are kept on lifelong daily aspirin. The optimal duration of anticoagulant use in NVAF patients who undergo LAAC for AIS-despite-OAC, is hotly debated. Some experts argue that these high embolic risk patients should remain on long-term OAC after LAAC in order to reduce recurrent embolic stroke risk. The LAAOS III study showed a 33% reduction in AIS risk when surgical LAAC was performed in addition to long-term OAC use in a patient population with NVAF who underwent cardiac surgery.^22^ Although a different surgical patient population, LAAOS III provided proof of concept that the combination of LAAC and long-term OAC is superior to OAC-only approach. In our study, 24.1% of NVAF patients who had LAAC after AIS-despite-OAC were kept on OAC at one year after the procedure. The rate of AIS recurrence was low at an average of 1.75 years follow-up and only 2 patients were lost to follow-up before 12 months. The recent FDA-required study, Primary Outcome Evaluation of a Next-Generation Left Atrial Appendage Closure Device (PINNACLE FLX) that resulted in approval of Watchman-FLX device showed that the majority of embolic events occur within the first year after LAAC, a finding in line with other published data.^23^ Among NVAF patients who sustained one or more embolic AIS-despite-OAC, our study shows low AIS rate despite conservative use of OAC after LAAC, over a relatively long follow-up compared to the other case series (**Table 4**).

**Table 4.**
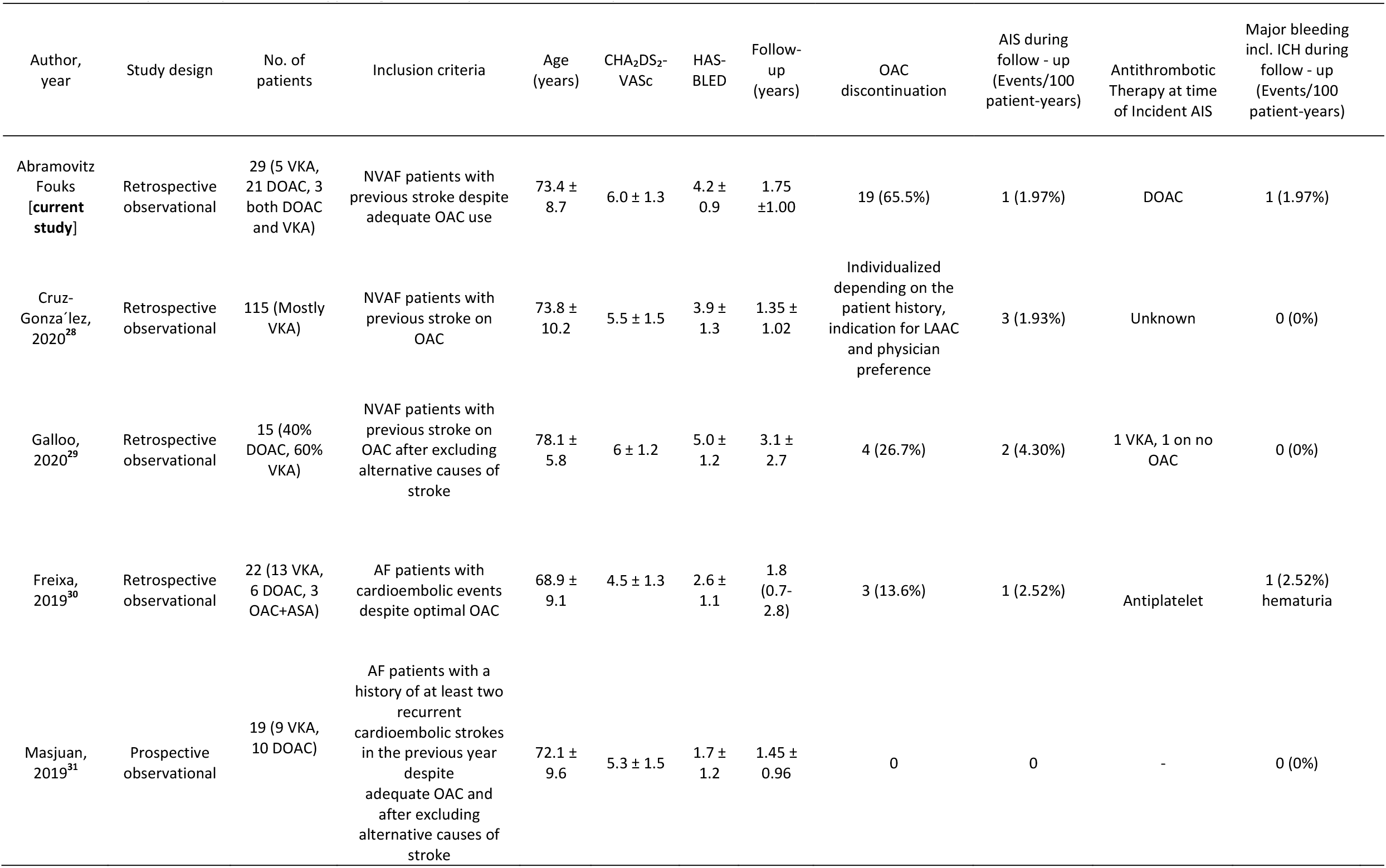
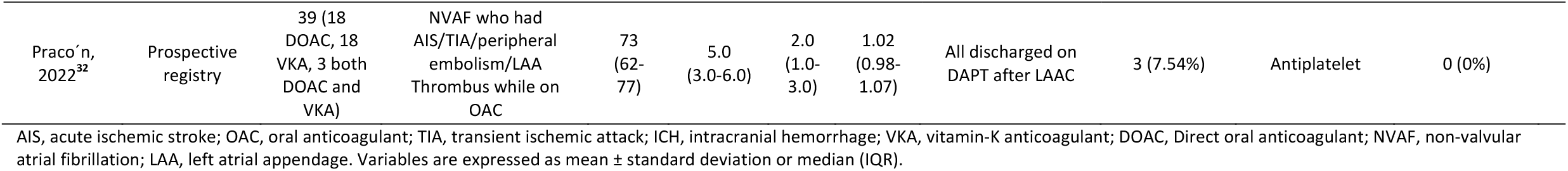
The efficacy and safety of Left atrial appendage closure in patients with AIS-despite-OAC.

The only patient who had an ICH in follow-up was on DOAC and aspirin therapy, 21.9 months after LAAC. Anti-thrombotic treatment after LAAC might be a challenging decision especially in high ischemic risk NVAF patients who also carry a high hemorrhagic risk. The patient who experienced ICH in follow-up had mixed location (deep and lobar) cerebral microbleeds on brain MRIs at the time of past AIS-despite-OAC. Mixed location cerebral microbleeds typically represent a more severe form of hypertensive cSVD.^24,25^ Such patients might undergo LAAC to be able to discontinue OAC in the absence of past AIS-despite-OAC. This patient was kept on DOAC and aspirin because of history of 4 AIS-despite-OAC. Post LAAC antithrombotic regimens varied in our cohort and such variability has been the rule rather than exception in previous reports as well.^26^ Among patients who received Watchman for LAAC between 2016 and 2018 included in the large LAAC Registry of the National Cardiovascular Data Registry, only 12.2% received the full post procedure antithrombotic treatment protocol studied in pivotal trials.^27^ As the question about the optimal antithrombotic treatment after LAAC in AIS-despite-OAC patients remains unanswered, assessing the individualized ischemic and hemorrhagic stroke risks for each patient might be the right approach until we have data from randomized trials. The main limitations of our study include its retrospective, observational nature, and the relatively small sample size. Despite these potential weaknesses, 93% of our patients had thorough follow-up for at least 1 year with a mean duration of 1.75 years follow-up. The current study also provides a high level of relevant detail for AIS-despite-OAC including detailed imaging review. Our sample size is average when compared to the other studies reported in **Table 4** but it should be remembered that LAAC is uncommonly performed after AIS-despite-OAC. Very detailed patient review and inclusion of patients who had LAAC specifically for AIS-despite-OAC are strengths of our study. About half of the patients had more than one AIS-despite-OAC and all were compliant with OAC use. Few of our patients had other potential etiologies for AIS (MGUS, heart failure, hemochromatosis, cSVD) but this would only elevate recurrent AIS risk and further emphasizes the success of LAAC as reflected by the low incidence AIS rates. A selection bias of patients who could tolerate the LAAC procedure could have influenced our results, but from an embolic and hemorrhagic prospective, the mean CHA_2_DS_2_-VASc and HAS-BLED of 5.96 and 4.24 respectively, represent a very high risk population who could reasonably be compared to previous studies which included patients with even lower risk scores (**Table 3**). Although we did not have a control group without LAAC, we compared our results to the expected AIS rates based on CHA_2_DS_2_-VASc scores of our own patients. We also reported relevant data from the previously published studies of AIS-despite-OAC who were maintained on anticoagulant therapy without LAAC as shown in Figure 1.

Our results show that the primary outcome of AIS after LAAC in a high embolic risk NVAF population who had AIS-despite-OAC was lower (1.97 per 100 patient-years) than the expected AIS rates calculated based on our cohort’s CHA_2_DS_2_-VASc scores (8.44 per 100 patient-years) and it was also lower than the rates previously reported in multiple studies that included patients who were kept on OAC as stroke prevention method (5.3-8.9 per 100 patient-years) as shown in Figure 1. LAAC might be beneficial to this population along with personalized post-LAAC antithrombotic treatment. RCTs are needed to confirm whether LAAC is a superior treatment approach for NVAF patients who had AIS-despite-OAC and identify the optimal post-LAAC antithrombotic regimen in this population.

## Non-standard Abbreviations and Acronyms

OAC: oral anticoagulants
NVAF: non-valvular atrial fibrillation
VKA: vitamin-K antagonists
AIS: acute ischemic stroke
DOAC: direct oral anticoagulant
ICH: intracerebral hemorrhage
LAAC: left atrial appendage closure
AF: atrial fibrillation
cSVD: cerebral small vessel disease
TEE: transesophageal echocardiogram
IR: incidence rate
RCT: randomized control trial

## References

1. Kleindorfer DO, Towfighi A, Chaturvedi S, Cockroft KM, Gutierrez J, Lombardi-Hill D, Kamel H, Kernan WN, Kittner SJ, Leira EC, et al. 2021 Guideline for the Prevention of Stroke in Patients With Stroke and Transient Ischemic Attack: A Guideline From the American Heart Association/American Stroke Association. Stroke. 2021;52. doi: 10.1161/str.0000000000000375

2. Stroke Prevention in Atrial Fibrillation Study. Final results. Circulation. 1991;84:527–539. doi: 10.1161/01.cir.84.2.527

3. Granger CB, Alexander JH, McMurray JJ, Lopes RD, Hylek EM, Hanna M, Al-Khalidi HR, Ansell J, Atar D, Avezum A, et al. Apixaban versus warfarin in patients with atrial fibrillation. N Engl J Med. 2011;365:981–992. doi: 10.1056/NEJMoa1107039

4. Connolly SJ, Ezekowitz MD, Yusuf S, Eikelboom J, Oldgren J, Parekh A, Pogue J, Reilly PA, Themeles E, Varrone J, et al. Dabigatran versus warfarin in patients with atrial fibrillation. N Engl J Med. 2009;361:1139–1151. doi: 10.1056/NEJMoa0905561

5. Patel MR, Mahaffey KW, Garg J, Pan G, Singer DE, Hacke W, Breithardt G, Halperin JL, Hankey GJ, Piccini JP, et al. Rivaroxaban versus warfarin in nonvalvular atrial fibrillation. N Engl J Med. 2011;365:883–891. doi: 10.1056/NEJMoa1009638

6. Giugliano RP, Ruff CT, Braunwald E, Murphy SA, Wiviott SD, Halperin JL, Waldo AL, Ezekowitz MD, Weitz JI, Spinar J, et al. Edoxaban versus warfarin in patients with atrial fibrillation. N Engl J Med. 2013;369:2093–2104. doi: 10.1056/NEJMoa1310907

7. Ruff CT, Giugliano RP, Braunwald E, Hoffman EB, Deenadayalu N, Ezekowitz MD, Camm AJ, Weitz JI, Lewis BS, Parkhomenko A, et al. Comparison of the efficacy and safety of new oral anticoagulants with warfarin in patients with atrial fibrillation: a meta-analysis of randomised trials. Lancet. 2014;383:955–962. doi: 10.1016/S0140-6736(13)62343-0

8. Gokcal E, Pasi M, Fisher M, Gurol ME. Atrial Fibrillation for the Neurologist: Preventing both Ischemic and Hemorrhagic Strokes. Curr Neurol Neurosci Rep. 2018;18:6. doi: 10.1007/s11910-018-0813-y

9. Seiffge DJ, De Marchis GM, Koga M, Paciaroni M, Wilson D, Cappellari M, Macha Md K, Tsivgoulis G, Ambler G, Arihiro S, et al. Ischemic Stroke despite Oral Anticoagulant Therapy in Patients with Atrial Fibrillation. Ann Neurol. 2020. doi: 10.1002/ana.25700

10. January CT, Wann LS, Calkins H, Chen LY, Cigarroa JE, Cleveland JC, Jr., Ellinor PT, Ezekowitz MD, Field ME, Furie KL, et al. 2019 AHA/ACC/HRS Focused Update of the 2014 AHA/ACC/HRS Guideline for the Management of Patients With Atrial Fibrillation: A Report of the American College of Cardiology/American Heart Association Task Force on Clinical Practice Guidelines and the Heart Rhythm Society in Collaboration With the Society of Thoracic Surgeons. Circulation. 2019;140:e125–e151. doi: 10.1161/CIR.0000000000000665

11. Gurol ME. Nonpharmacological Management of Atrial Fibrillation in Patients at High Intracranial Hemorrhage Risk. Stroke. 2018;49:247–254. doi: 10.1161/STROKEAHA.117.017081

12. Kang DW, Chalela JA, Ezzeddine MA, Warach S. Association of ischemic lesion patterns on early diffusion-weighted imaging with TOAST stroke subtypes. Arch Neurol. 2003;60:1730–1734. doi: 10.1001/archneur.60.12.1730

13. Regenhardt RW, Das AS, Ohtomo R, Lo EH, Ayata C, Gurol ME. Pathophysiology of Lacunar Stroke: History’s Mysteries and Modern Interpretations. J Stroke Cerebrovasc Dis. 2019;28:2079–2097. doi: 10.1016/j.jstrokecerebrovasdis.2019.05.006

14. Das AS, Regenhardt RW, Feske SK, Gurol ME. Treatment Approaches to Lacunar Stroke. J Stroke Cerebrovasc Dis. 2019;28:2055–2078. doi: 10.1016/j.jstrokecerebrovasdis.2019.05.004

15. Benz AP, Hohnloser SH, Eikelboom JW, Carnicelli AP, Giugliano RP, Granger CB, Harrington J, Hijazi Z, Morrow DA, Patel MR, et al. Outcomes of patients with atrial fibrillation and ischemic stroke while on oral anticoagulation. Eur Heart J. 2023. doi: 10.1093/eurheartj/ehad200

16. Tanaka K, Koga M, Lee KJ, Kim BJ, Park EL, Lee J, Mizoguchi T, Yoshimura S, Cha JK, Lee BC, et al. Atrial Fibrillation-Associated Ischemic Stroke Patients With Prior Anticoagulation Have Higher Risk for Recurrent Stroke. Stroke. 2020;51:1150–1157. doi: 10.1161/STROKEAHA.119.027275

17. Yaghi S, Henninger N, Giles JA, Leon Guerrero C, Mistry E, Liberman AL, Asad D, Liu A, Nagy M, Kaushal A, et al. Ischaemic stroke on anticoagulation therapy and early recurrence in acute cardioembolic stroke: the IAC study. J Neurol Neurosurg Psychiatry. 2021;92:1062–1067. doi: 10.1136/jnnp-2021-326166

18. Paciaroni M, Caso V, Agnelli G, Mosconi MG, Giustozzi M, Seiffge DJ, Engelter ST, Lyrer P, Polymeris AA, Kriemler L, et al. Recurrent Ischemic Stroke and Bleeding in Patients With Atrial Fibrillation Who Suffered an Acute Stroke While on Treatment With Nonvitamin K Antagonist Oral Anticoagulants: The RENO-EXTEND Study. Stroke. 2022;53:2620–2627. doi: 10.1161/STROKEAHA.121.038239

19. Tokunaga K, Koga M, Itabashi R, Yamagami H, Todo K, Yoshimura S, Kimura K, Sato S, Terasaki T, Inoue M, et al. Prior Anticoagulation and Short-or Long-Term Clinical Outcomes in Ischemic Stroke or Transient Ischemic Attack Patients With Nonvalvular Atrial Fibrillation. J Am Heart Assoc. 2019;8:e010593. doi: 10.1161/JAHA.118.010593

20. Polymeris AA, Meinel TR, Oehler H, Holscher K, Zietz A, Scheitz JF, Nolte CH, Stretz C, Yaghi S, Stoll S, et al. Aetiology, secondary prevention strategies and outcomes of ischaemic stroke despite oral anticoagulant therapy in patients with atrial fibrillation. J Neurol Neurosurg Psychiatry. 2022;93:588–598. doi: 10.1136/jnnp-2021-328391

21. Kristinsson SY, Pfeiffer RM, Bjorkholm M, Goldin LR, Schulman S, Blimark C, Mellqvist UH, Wahlin A, Turesson I, Landgren O. Arterial and venous thrombosis in monoclonal gammopathy of undetermined significance and multiple myeloma: a population-based study. Blood. 2010;115:4991–4998. doi: 10.1182/blood-2009-11-252072

22. Whitlock RP, Belley-Cote EP, Paparella D, Healey JS, Brady K, Sharma M, Reents W, Budera P, Baddour AJ, Fila P, et al. Left Atrial Appendage Occlusion during Cardiac Surgery to Prevent Stroke. N Engl J Med. 2021;384:2081–2091. doi: 10.1056/NEJMoa2101897

23. Doshi SK, Kar S, Sadhu A, Horton R, Osorio J, Ellis C, Stone J, Jr., Shah M, Dukkipati SR, Adler S, et al. Two-Year Outcomes With a Next-Generation Left Atrial Appendage Device: Final Results of the PINNACLE FLX Trial. J Am Heart Assoc. 2023;12:e026295. doi: 10.1161/JAHA.122.026295

24. Pasi M, Charidimou A, Boulouis G, Auriel E, Ayres A, Schwab KM, Goldstein JN, Rosand J, Viswanathan A, Pantoni L, et al. Mixed-location cerebral hemorrhage/microbleeds: Underlying microangiopathy and recurrence risk. Neurology. 2018;90:e119–e126. doi: 10.1212/WNL.0000000000004797

25. Tsai HH, Pasi M, Tsai LK, Chen YF, Lee BC, Tang SC, Fotiadis P, Huang CY, Yen RF, Jeng JS, et al. Microangiopathy underlying mixed-location intracerebral hemorrhages/microbleeds: A PiB-PET study. Neurology. 2019;92:e774–e781. doi: 10.1212/WNL.0000000000006953

26. Cohen JA, Heist EK, Galvin J, Lee H, Johnson M, Fitzsimons M, Slattery K, Ghoshhajra B, Sakhuja R, Ha G, et al. A comparison of postprocedural anticoagulation in high-risk patients undergoing WATCHMAN device implantation. Pacing Clin Electrophysiol. 2019;42:1304–1309. doi: 10.1111/pace.13796

27. Freeman JV, Higgins AY, Wang Y, Du C, Friedman DJ, Daimee UA, Minges KE, Pereira L, Goldsweig AM, Price MJ, et al. Antithrombotic Therapy After Left Atrial Appendage Occlusion in Patients With Atrial Fibrillation. J Am Coll Cardiol. 2022;79:1785–1798. doi: 10.1016/j.jacc.2022.02.047

28. Cruz-Gonzalez I, Gonzalez-Ferreiro R, Freixa X, Gafoor S, Shakir S, Omran H, Berti S, Santoro G, Kefer J, Landmesser U, et al. Left atrial appendage occlusion for stroke despite oral anticoagulation (resistant stroke). Results from the Amplatzer Cardiac Plug registry. Rev Esp Cardiol (Engl Ed). 2020;73:28–34. doi: 10.1016/j.rec.2019.02.013

29. Galloo X, Carmeliet T, Prihadi EA, Lochy S, Scott B, Verheye S, Schoors D, Vermeersch P. Left atrial appendage occlusion in recurrent ischaemic stroke, a multicentre experience. Acta Clin Belg. 2022;77:255–260. doi: 10.1080/17843286.2020.1821494

30. Freixa X, Cruz-Gonzalez I, Regueiro A, Nombela-Franco L, Estevez-Loureiro R, Ruiz-Salmeron R, Bethencourt A, Gutierrez-Garcia H, Fernandez-Diaz JA, Moreno-Samos JC, et al. Left Atrial Appendage Occlusion as Adjunctive Therapy to Anticoagulation for Stroke Recurrence. J Invasive Cardiol. 2019;31:212–216.

31. Masjuan J, Salido L, DeFelipe A, Hernandez-Antolin R, Fernandez-Golfin C, Cruz-Culebras A, Matute C, Vera R, Perez-Torre P, Zamorano JL. Oral anticoagulation and left atrial appendage closure: a new strategy for recurrent cardioembolic stroke. Eur J Neurol. 2019;26:816–820. doi: 10.1111/ene.13894

32. Pracon R, Zielinski K, Bangalore S, Konka M, Kruk M, Kepka C, Trochimiuk P, Debski M, Przyluski J, Kaczmarska E, et al. Residual stroke risk after left atrial appendage closure in patients with prior oral anticoagulation failure. Int J Cardiol. 2022;354:17–21. doi: 10.1016/j.ijcard.2022.02.030

